# “Assessment of Referral System on Maternal Services in Cagayan De Oro City”

**DOI:** 10.1101/2022.03.31.22273250

**Authors:** Arbeen Acosta Laurito

**Affiliations:** Justiniano R. Borja General Hospital

**Keywords:** compliance, forms utilization, maternal care services, referral system, standard referral form, women of reproductive age

## Abstract

A retrospective document analysis study aimed to assess the referral system on maternal services based on the referral forms of referred women of reproductive age from the referral facilities to the receiving hospital from January to December 2019. The specific objectives of the study were to describe the use of standard referral forms, compliance of the healthcare workers in using the standard form based on 14 criteria, and the utilization of relevant data related to maternal services based on 16 criteria. There were 3330 referral forms received by the receiving facility on different formats of forms during the study period. A random sampling of 384 referral forms was used as study population. Among 384 referral forms (random sampling), only 126 (31.8%) used the standard referral forms. The compliance of these referral forms using 14 criteria showed that 116 (92.06%) referral forms complied with only 51-75% of the criteria, and none of the referral forms complied with all the 14 criteria. On assessing the data entries among 384 referral forms with different formats, there were six data entries consistently used more than 60% by the healthcare providers which were not part of the printed form: last menstrual period (67.87%), expected date of confinement (64.84%), fundic height (63.04%), fetal heart beat (60.76%), birthweight (62.59%), and age of gestation (60%). Based on the 16 criteria, majority of referral forms (210) utilized 51-75% of the 16 criteria, 122 referral forms utilized 76-99%, 51 forms utilized 26-50%, and 1 form utilized less than 25% of the data entries. Several studies documented that referral forms and functional referral systems are vital to an improved maternal mortality rate (MMR) and infant mortality rate (IMR). Therefore, as part of a continued quality referral system, it is highly recommended that the required referral forms be re-assessed, revised, and regularly monitored on its form compliance and utilization.

## Introduction

Improving referral system in maternal services means improving maternal mortality rate (MMR) and infant mortality rate (IMR) (DOH AO No. 2016-0035). Studies documented that quality referral system is a component in health program implementation and in providing quality healthcare services (Davidow et al. 2018; Blake-Lamb et al. 2016). The Philippine government is challenged and responsive to these indicators by implementing the Philippine Health Agenda in line with the Sustainable Development Goals to access quality health services made available in different levels of care through Service Delivery Networks (SDN) (DOH AO No. 2017-0014).

Since the referral system is a process which delivers a continuum of maternal (and neonatal) services, the two-way process is under-estimated, and should functioned as a ‘referral loop” (Murray and Pearson, 2005), which should involve the public, private, community-based health care providers and health care facilities, which can take the form of a vertical, horizontal, or diagonal relationships (DOH-NMHRS, 2015).

Various studies strengthened the continuum of care (Okawa et al. 2019; Kerber et al. 2007). In Cambodia, Kikuchi et al (2018) assessed the completion rate of the continuum of care and examined the factors associated with it. The spectrum of continuum of care of pregnant women consisted of the following: antenatal care (4 visits), delivery by skilled birth attendant, facility-based delivery, and postnatal care (2 visits). Consolidated Philippine data comparing 2015 FHS and 2018 FHSIS showed that skilled birth attendant (80% to 94.18%) and facility-based delivery (80% to 92.59%) significantly improved (DOH, 2018). But there is a great need to re-focus and improved both antenatal care (from 75% to 52.55%) and postnatal care (from 80% to 57.43%).

Referring to the manual on Northern Mindanao Health Referral System (DOH-NMHRS, 2015), referral loop consists mainly of the referring or initiating health care provider or health facility which prepares an outward referral to communicate the patient’s condition, status and pre-referral interventions provided, and the reasons for referral; receiving health care provider or health facility is the one who accepts the referred case, provides the needed care, and prepares the back referral at the end of their health care involvement, to let the initiating facility know what has been done. The full-service implementation of back-referral to complete the loop referral is essential at this stage and will serve as a feedback mechanism to the referring facility.

Regionally, the Department of Health, responded to improve the referral system by drafting NMHRS, institutionalized the local usage of proforma, patterned from the WHO (2015) format (Appendix A), which was also followed by the DOH Manual on Health Referral System of 2002. A local study by Adap et al. (2017), evaluated the health referral system in Cagayan De Oro City based on the availability of document retrieved per facilities active in the public health referral system. Deficiencies of consolidated records, referral logbooks, registry and reporting were noted in some health facilities.

### Statement of the Problem

**T**his study aims to assess the referral system for maternal services in Cagayan De Oro City. Specifically, it deals with the following:

1. To describe and assess the referred women of reproductive age using standard Referral Form.
2. To assess the compliance of the health care professionals on the use of standard referral forms using the 14 criteria.
3. To assess the utilization of the referral forms based on 16 criteria for maternal services

### Conceptual Framework

This study was grounded from the conceptual framework for Referral Care for Maternal, Neonatal and Child Health and Nutrition-Family Planning (MNCHN-FP) Services (DOH-JHPIEGO, 2016). In order to conceptualized the two-way process and to ensure the continuum of care, the referral system process involves the public, private, community-based health care providers and health care facilities, which can take the form of a vertical, horizontal, or diagonal relationships.

### Scope and Limitation

The study population were limited to the women of reproductive aged (WRA, 15– 49 years old) who were referred from their respective barangay health centers (referring facility) of Cagayan De Oro City to avail higher level of maternal services. Referral forms were retrieved and data collection were gathered retrospectively based on these forms. There were no interactions nor data verification with the teenage and women of reproductive age. Referral forms were randomly selected among the compiled forms at JRBGH from January 2019 to December 2019. Referred WRA due to gynecologic cases, non-residents of Cagayan De Oro City, resident of the city but referred from other health facilities outside of Cagayan De Oro City, and use of prescription form as Referral form were excluded from the study.

### Definition of Terms

#### 1. Compliant Referal Form

Refers to the availability of 14 data entries filled up in the Referral form and detached Acknowledgement Receipt based on the standard Referral form (14/14, 100%). Moreover, it was based on the following 14 criteria: (1) name of the health care provider referring, (2) referring facility, (3) accompanied by the healthcare provider, (4) date and (5) time of referral, (6) age and (7) address, (8) main reason for referral, (9) major findings, (10) treatment given, (11) treatment before and (12) during referral, and (13) information given to the WRA or companion. An attached or a detached (14) Acknowledgement Receipt were considered as the last criteria as presented in the DOH-NMHRS 2015 (See Appendix B).

#### 2. Non-Compliant Referral Form

Refers to the score of 13 criteria and below based on the standard Referral form Non-compliant Referral forms were categorized as follows; 0-25% compliant (0-3 entries), 26-50% compliant (4-7 entries), 51-75% compliant (8-10 entries), and 76-99% compliant (11-13 entries).

#### 3. Complete Referral Form

Refers to the received Referral forms (in any format, eg. Clinical Referral Form [Appendix C]) with 16 criteria completely filled up and an attached document (16/16, 100%), which are necessary for the continuity of maternal services. These criteria are listed in Appendix D, as follows; 1) age of the patient, 2) blood pressure monitoring, 3) OB score written as G_P_, 4) reason for referral, 5) date of referral, 6) name of Referring facility, 7) name of the healthcare personnel from the referring unit, 8) date received, 9) detached acknowledgement receipt, (10) last menstrual cycle (LMP), (11) expected date of confinement (EDC), (12) age of gestation (AOG), (13) fundic height (FH), (14) fetal heart beat (FHB), (15) body weight of WRA (BW), and (16) use of partograph.

#### 4. In-complete Referral Forms

Refers to the 15 criteria and below for maternal services among the received Referral forms. These were categorized, as follows; 0-25% complete (0-4 entries), 26-50% complete (5-8 entries), 51-75% complete (9-12 entries), and 76-99% complete (13-15 entries).

#### 5. Standard Referral Form

Refers to the Referral Form published by DOH-Northern Mindanao Health Referral System Manual, 2nd ed. (2015) labeled as Maternal/ BEmONC Referral Form. This form was adopted from WHO (2003). It has 2 sections which were used for referring both WRA and the newborn delivered by the WRA (Maternal and Baby sections) using the same Referral form. Acknowledgement Receipt is located at the bottom of the form. This was filled-up, detached and given to the accompanying staff or driver who brought the WRA to the receiving facility; and documented by the referring facility to attest that WRA was received by the receiving facility.

### Research Design

This is a retrospective-document analysis designed to assess the referral system among randomly selected referred women of reproductive aged (WRA) 15 to 49 years old in Cagayan De Oro City from January 2019 to December 2019 to the receiving facility (LGU-owned level 1 public hospital).

### Sampling Procedure

There were 3,330 WRA referrals from the referring facilities of Cagayan De Oro City to the receiving facility for 2019. A total of 384 study population was selected through random sampling for every month with 32 samples per month from January 2019 to December 2019 **(Figure 1.)** This is to meet the minimum sample to achieve 95% level of confidence, maximum tolerance of 5% and an estimated population of 50%.

**Figure 1.**
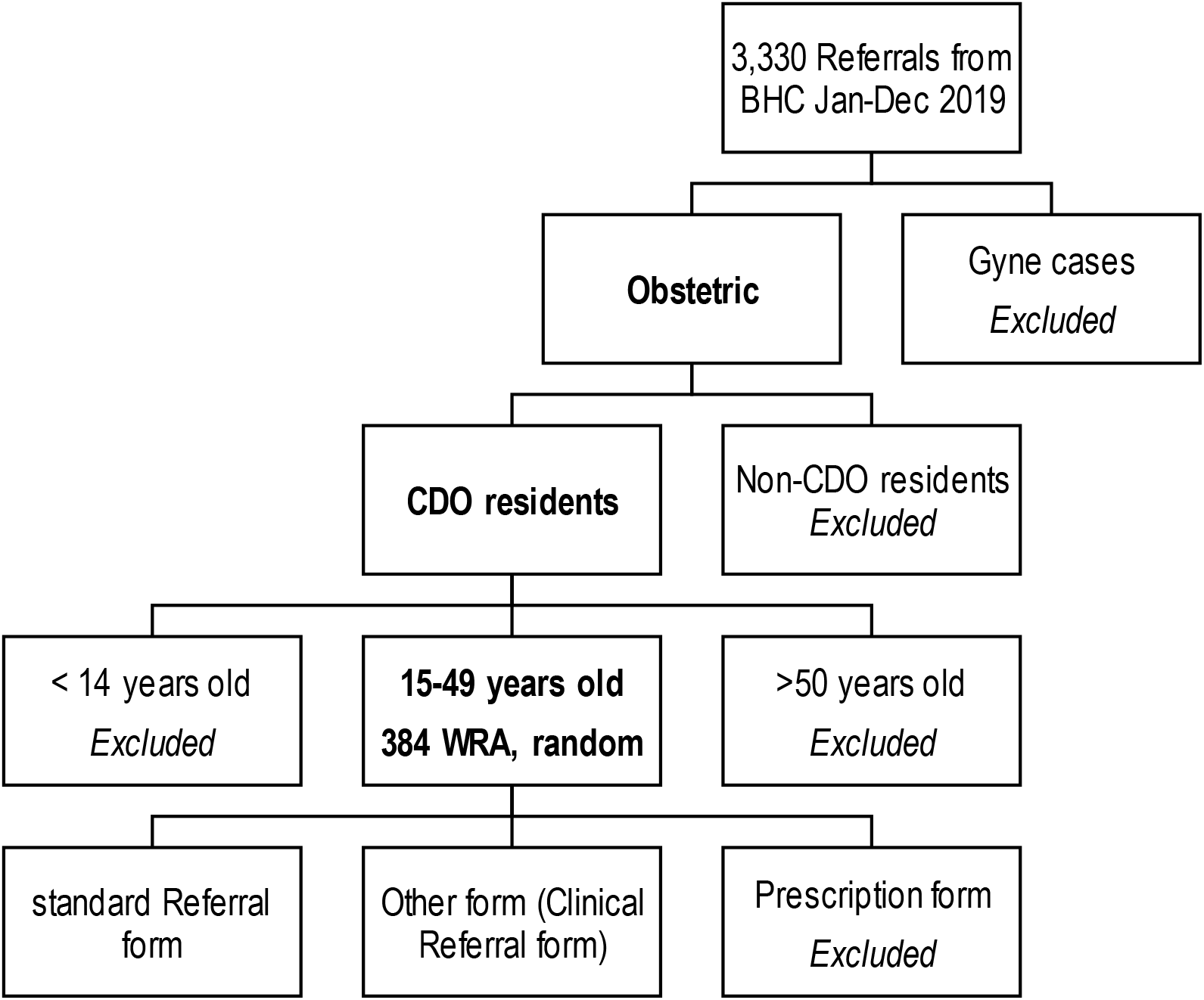
Random sampling of the study population on referral forms among women of reproductive age referred from the referring facilities, January-December 2019.

### Statistical Instruments/ Procedure

Descriptive statistics were used such as frequency, mean and percentages to present collected primary data representing the characteristics of the referral system in Cagayan De Oro City and the secondary data from the compiled Referral forms by the receiving facility. Quality of the randomly selected Referral forms were assessed based on the minimum elements of a standard Referral form as required by WHO 2003 (adopted by DOH-NMHRS, 2015). Compliant Referral forms refer to a score of 14/14 based on 14 criteria from then standard or 100%, and Referral forms utilization refers to a score of 16/16 or 100% from the criteria on maternal services (eg. LMP, EDC).

In comparison with the standard Referral form, the compiled Referral forms assessed as compliant (100%, 14/14 criteria) based on the entries filled up specifically on the boxes for Woman column (left side of the form), and the presence or detached Acknowledgement Receipt (Appendix B). Referral forms other than the standard referral form (eg. prescription form) were not included. Based on the 14 criteria (compared to the standard Referral form), non-compliant Referral forms were categorized as follows; 0-25% compliant (0-4 entries), 26-50% compliant (5-7 entries), 51-75% compliant (8-10 entries), and 76-99% compliant (11-13 entries).

These 13 entries were the following: (1) name of the health care provider referring, (2) referring facility, (3) accompanied by the healthcare provider, (4) date and (5) time of referral, (6) age and (7) address, (8) main reason for referral, (9) major findings, (10) treatment given, (11) treatment before and (12) during referral, and (13) information given to the WRA or companion. The presence or a detached (14) Acknowledgement Receipt was the last criteria (Appendix B).

Moreover, Referral forms using standard Referral form and other forms (eg. Clinical Referral Form) were assessed for the relevance of data with specific criteria on maternal services (16/16, 100%). Fifteen (15) criteria using the form and the use of partograph were the variables for maternal services. Likewise, using the 16 criteria for maternal services, Referral forms were categorized accordingly; 0-25% utilized the 16 criteria (0-4 entries), 26-50% utilized (5-8 entries), 51-75% utilized (9-12 entries), and 76-99% utilized (13-15 entries). Frequency, percentages and means were used for presenting the data. These 16 variables are necessary for the re-evaluation of WRA especially if in active labor, shortens the history taking, and more focused on their needs. These variables were presented in the Referral Form Assessment Tool (Maternal services) [Appendix D], such as: 1) age of the patient, 2) blood pressure monitoring, 3) OB score written as G_P_, 4) reason/s for referral, 5) date of referral, 6) name of referring facility, 7) name of the health care provider from the referring unit, 8) date received, 9) detached acknowledgement receipt, (10) last menstrual cycle (LMP), expected date of confinement (EDC), (12) age of gestation (AOG), (13) fundic height (FH), (14) fetal heart beat (FHB), (15) body weight of WRA (BW), and (16) use of partograph.

Appropriate correlations among variables and between the data collected from the referring facilities and receiving facility were analyzed qualitatively.

## RESULTS AND DISCUSSIONS

### A. Profile of referred women of reproductive age

Census from the LGU-owned level 1 hospital covering the months of January 2019 to December 2019 showed 3,330 referred WRA at the receiving facility or 52% of the 6,456 deliveries for 2019. The peak month of referrals was on June (413, 12.4%) and the least number of referred WRA was on December 2019 (195, 5.86%). The receiving facility received an average of 278 referred WRA per month from the 80 barangays of Cagayan de Oro City and neighboring municipalities of Bukidnon (447) and Misamis Oriental (253).

Majority of these 3330 referred WRA age range was the young adult at 20-24 yr old (907), followed by the 25-29 yr old (753), teenagers 15-19 yr old (668), 30-34 yr old (504), 35-39 yr old (277), and 40-44 yr old (132). Beyond the age of WRA, there were 12 women aged 14 yr old and below while 15 women were referred with the age 45 yr old and above. The youngest woman referred was 13 years old while the oldest was 56 years old. Despite its importance in filling up the Referral form for age, there were 62 Referral forms with no age entry.

Among the 3330 reasons for referral, it was broadly classified as maternal-related reasons (1862, 55.92%), non-maternal related (751, 22.55%), and unfilled reasons for referral (717, 21.53%). Maternal-related reasons for referrals were sub-classified into high risk (1301), low risk (222), no risk at all (339).

There were 1301 WRA referred due to maternal high-risk indicators, as follows; teenage pregnancy (383), elevated blood pressure (143), anemia/ low Hemoglobin (115), previous Cesarian section (90), and breech presentation (88).

These reasons for referrals categorized as low risk had 222 WRA referred from referring facilities included; hepatitis B, reactive (38), previous history of abortion (32), maternal medical problems (22), urinary tract infection (19), and fundic height of variable measurements (17).

Majority of these referred WRA came in by themselves without accompanying health care providers from the referring facilities. And it was observed that Acknowledgement receipts were still attached (2,556, 76.76%) to the compiled Referral forms and only 727 (21.83%) were detached which followed the protocol. About 47 (1.41%) Referral forms had no Acknowledgement receipts since they were not in the standard Referral forms (eg. prescription form).

Referred WRA were initially seen by health care providers at the referring facilities. These women of reproductive age were seen and referred mostly by midwives who specifically handled deliveries for WRA (1033, 31.02%), followed by medical doctors (282, 8.47%), 129 by registered nurses (3.87%), and 5 WRA by barangay nutrition scholars themselves (BNS, 0.15%). However, there were 401 (12.04%) Referral forms with no names for health care providers. The distribution of health care providers by professions as mentioned above maybe changed if the 1480 names (only) of health care providers are appropriately re-classified, but it is beyond the scope of the study.

Utilization Relevant and related data to maternal services were described in Figure 1. They were not part of the printed form of the standard Referral form. And all of these data were handwritten or filled up more than 60% of all Referral forms received. Thus, these data were necessary for the initial assessment of WRA, early determination of impending complications, appropriate management and continuity of care at the level of the receiving facility. Therefore, these data are highly recommended for inclusion and be utilized in the revised form template whenever WRA will be referred to other health facilities.

### B. Profile of the used Referral Forms based of the standard BEmNOC/CEmONC Referral Form

To assessed the compliance of the Referral forms from the referring facilities, only 14 criteria were used in the study since other 4 data entry were never filled up. The following 14 criteria were enumerated on **Table 1** with its corresponding frequencies and percentages. The filled-up Acknowledgement receipts (14) whether attached and or detached was the last criteria (14/14, 100%).

**Table 1.** Frequency and percentage distribution of data entries filled up in the standard referral forms. (n=126)

| Data Entries | Parts of the Referral Form | With entry |  | No entry |  |
| --- | --- | --- | --- | --- | --- |
|  |  | Frequency | % | Frequency | % |
| 1 | Name of the health care provider referring | 113 | 89.68 | 13 | 10.32 |
| 2 | Referring facility | 116 | 92.06 | 10 | 7.94 |
| 3 | Accompanied by the health care provider | 2 | 1.59 | 124 | 98.41 |
| 4 | Date of referral | 116 | 92.06 | 10 | 7.94 |
| 5 | Time of referral | 103 | 81.75 | 23 | 18.25 |
| 6 | Age of WRA | 125 | 100.00 | 1 | 0.79 |
| 7 | Address of WRA | 123 | 97.62 | 3 | 2.38 |
| 8 | Main reason for referral | 67 | 53.17 | 59 | 46.83 |
| 9 | Major findings | 125 | 99.21 | 1 | 0.79 |
| 10 | Treatment given | 28 | 22.22 | 98 | 77.78 |
| 11 | Treatment before | 26 | 20.63 | 100 | 79.37 |
| 12 | Treatment during referral | 2 | 1.59 | 124 | 98.41 |
| 13 | Information given to the WRA or companion | 114 | 90.48% | 12 | 9.52 |
| 14 | Attached or detached Acknowledgement Receipt | 40<br>(Detached) | 31.75 | 86<br>(Attached) | 68.25 |

**Table 2.** Frequency and percentage of data entries filled up in the referral forms according to relevance of data on maternal services. (n=384)

| Data entries | Parts of the Referral Form | With entry |  | No entry |  |
| --- | --- | --- | --- | --- | --- |
|  |  | Frequency | % | Frequency | % |
| 1 | Age of WRA | 380 | 98.96 | 4 | 1.04 |
| 2 | Blood pressure | 337 | 87.76 | 47 | 12.24 |
| 3 | OB score (G-P-) | 280 | 72.92 | 104 | 27.08 |
| 4 | Reason for referral | 319 | 83.07 | 65 | 16.93 |
| 5 | Date of referral | 363 | 94.53 | 21 | 5.47 |
| 6 | Name of referring facility | 276 | 71.88 | 108 | 28.12 |
| 7 | Name of health care personnel | 384 | 100 | 0 | 0 |
| 8 | Date received | 360 | 93.75 | 24 | 6.25 |
| 9 | Detached Acknowledgement receipt | 81 | 21.09 | 303 | 78.91 |
| 10 | Last menstrual cycle (LMP) | 261 | 67.87 | 123 | 32.13 |
| 11 | Expected date of confinement (EDC) | 249 | 64.84 | 135 | 35.16 |
| 12 | Age of gestation (AOG) | 230 | 60.00 | 154 | 39.99 |
| 13 | Fundic height (FH) | 242 | 63.04 | 142 | 36.96 |
| 14 | Fetal heart beat (FHB) | 233 | 60.76 | 151 | 39.24 |
| 15 | Body weight of WRA | 240 | 62.59 | 144 | 37.41 |
| 16 | Use of partograph | 8 | 2.20% | 356 | 97.8 |

Among the 384 randomly selected Referral forms, only 126 (31.8%) forms used the standard BEmONC/CEmONC Referral form or Maternity/BEmONC Referral form (Appendix B). From these 126 standard Referral forms, the compliance of the health care providers in filling up data was assessed based on the above-mentioned 14 criteria. Table 1 presented the 14 criteria with the frequency of entries or no entries in the criteria and its corresponding percentages.

The following discussions followed the order of entry in the standard Referral form. Under the REFERRAL RECORD section, the following are required; 1) Name of the staff who refer WRA, 2) Name of facility and entry on the 3) Accompanied by health worker.

Among health care staff who referred WRA (113), majority were seen and referred by the midwives (21) and only two were referred by the Medical Officers (handle 3-5 barangays in clusters). The other HCP indicated were their names only and the profession cannot be assessed. Among the referred WRA, they were 126 WRA who were referred from top 4 facilities (Barangays) among the 80 barangays, namely: Barangay Tablon (15), Lapasan (11), Camaman-an (11) and Bugo (8), and the City Health Office (8).

There were 66 maternal-related reasons for referral and these were; teenage pregnancy (19 cases), followed by elevated blood pressure (9), low hemoglobin count (6), breech presentation (6), ruptured or leaking bag of water (5), history of previous cesarian section (4), primigravida (or first pregnancy, 3 cases), and post-term (42 weeks of pregnancy/3 cases). There were 41 n**on-maternal related reason**s for referrals; for ultrasonography (3), vaginal spotting (3), non- maternal care package (MCP) accredited (5) facility, patient’s choice (6), and non-emergency cases (7), and the rest had 1 case each. There were 19 cases where reasons for referrals were not indicated.

Of these 126 Referral forms, majority of the required entries were filled up. However, majority of the required data entries had no entries such as; (3) Accompanied by the health care provider, (10) Treatment given, (11) Treatment before referral, and (12) treatment during referral.

Moreover, current study showed that among the 126 standard Referral forms, only 31.75% (40 forms) had detached Acknowledgement Receipts. These were presumed to be received by the referring facility and other circumstances (eg lost receipts). It is beyond the scope of the study to confirm these Acknowledgement Receipts at the referring facilities. And in addition, there were 68.25% (86 forms) found to have attached Acknowledgement Receipts. Among the Referral forms with an attached Acknowledgement Receipts, 48 (55.81%) had data filled up while 38 Referral forms (44.19%) had no entries at all.

### C. Compliance to the used Referral Forms based on 14 criteria

The compliance of the used Referral forms was based on the currently recommended DOH-NMHRS (2015) Manual with 13 filled up data entries in the standard Referral form plus the detached Acknowledgement Receipt with a score of 14/14 or 100% compliant. This study showed that all 126 Referral forms were non-compliant to the identified 14 criteria since no form had complied ALL of the 14 variables (**Figure 2)**.

**Figure 2.**
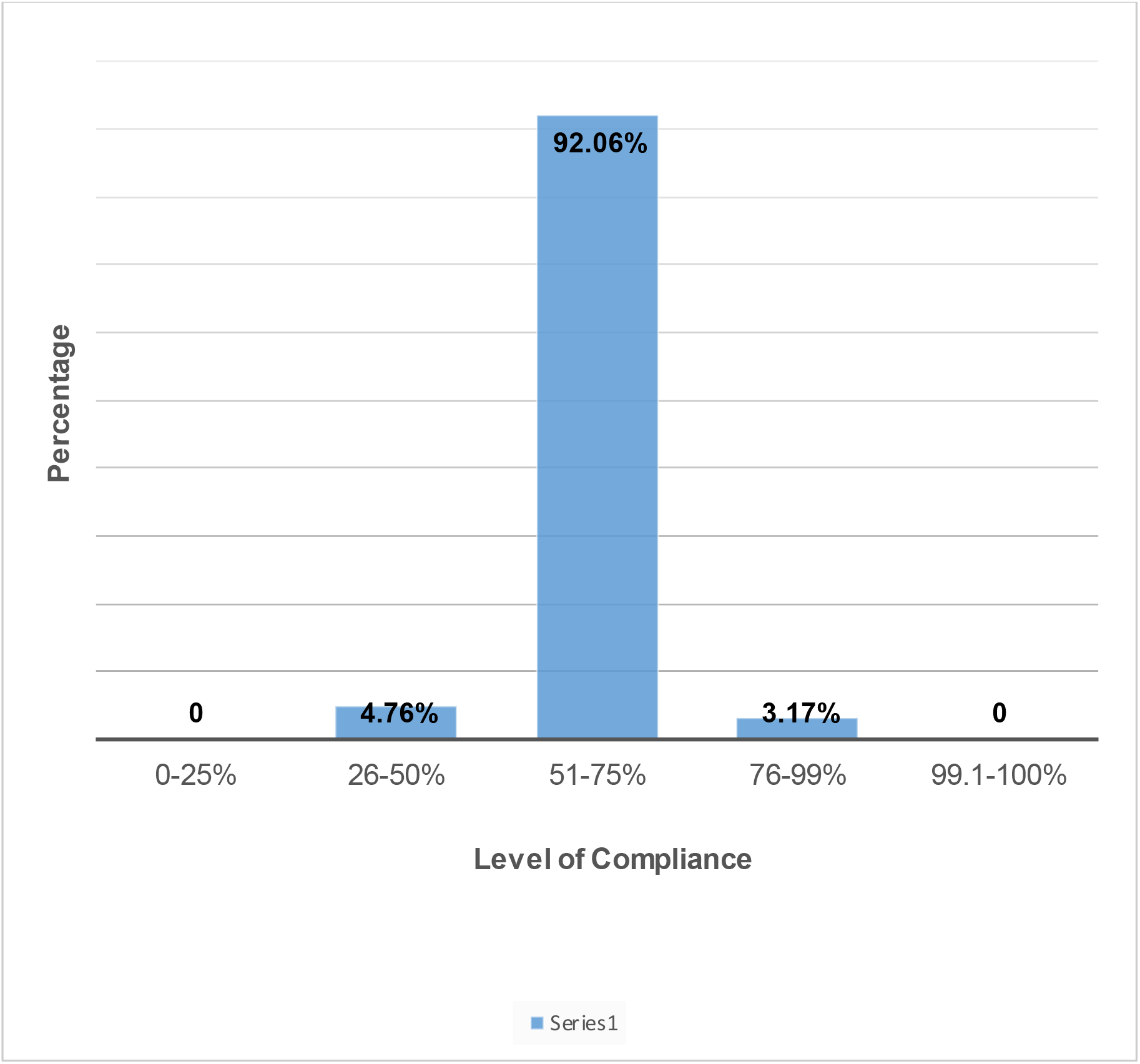
The distribution of referral forms based on its compliance (n=126).

### D. Assessment of the Referral forms on utilization of data related to maternal services based on 16 criteria

These WRA had their first contact with the health care system at the level of the barangay health centers during their antenatal care (ANC). WRA were referred to the receiving facility to avail the higher level of care of which referring facilities were not capable of handling complicated cases. And the intake of relevant high-quality data started at the level of the referring facility which were confirmed on WRA frequent visits at the referring facilities during ANC. Non-conformity and or deviations from these data would mean a complicated pregnancy and urgently required referral to a hospital facility. The following discussions summarized the importance of inclusion of these 16 criteria

The age of WRA is important because interventions in each age group varies. And based on general population, certain age group has more prevalence of pregnancy-related complications. The obstetric score (OB score) represented the pregnancy history of the WRA, abortion history and successful deliveries and make the receiving facility health care providers cautious of these previous complications. Other criteria (and its variations) included would mean complicated pregnancy, such as: initial blood pressure (elevated would mean hypertension-related pregnancy), last menstrual cycle (beyond 42 weeks means post-maturity), expected date of confinement (prepared the WRA for delivery, and post-maturity if beyond 42 weeks), age of gestation, fundic height (means a small or large baby or malpresentation), fetal heart beat (absence means death and needs urgent delivery and or resuscitation), body weight (maternal obesity may have complication with hypertension or delivery of large baby). The remaining 4 criteria were the demographics of WRA, in terms of; the reasons for referral, date referred and received, and name of health care personnel from the referring facility. Furthermore, the last two (2) criteria represented the importance of the role of both referring (using the partograph as recommended by the NMHRS protocol) and receiving facilities (giving of filled up Acknowledgement Receipt). Based on these 16 criteria, discussions below gave us the results from the randomly selected 384 Referral forms regardless of the form style.

In line with the presentation of the above 16 criteria, the researcher emphasized that these data were not included in the printed template of the different forms style of Referral forms, namely; OB score, (10) LMP, (11) EDC, (12) AOG, (13) FH, (14) FHB, and partograph use.

The six variables shown in **Figure 4**, were written by the health care providers even if no section in the referral from required them to do so. These data were also available in any section of the Referral form regardless of the form style. In addition, these variables were used consistently by more than 60% of the health care providers in the Referral forms except for the use of partograph. These variables are vital for the continuing care of the maternal services from referring up to the receiving health care facilities. The utilization of these variables was generally accepted and understood by majority of the HCP which were then utilized for the continuity of care at the level of the receiving facility. Any deviations from each of these variables would mean an urgent indication for a WRA referral to higher level of care for appropriate management.

### E. Utilization of relevant data on related to maternal services based on 16 criteria

The study had showed the utilization of 14 data entries in the Referral forms (regardless of the Referral form style), and the (15) detached Acknowledgement Receipt and an attached (16) partograph, making a score of 16/16 (100%). Figure 3 presented the percentage level of utilization of the randomly selected 384 referral forms from these WRA.

**Figure 3.**
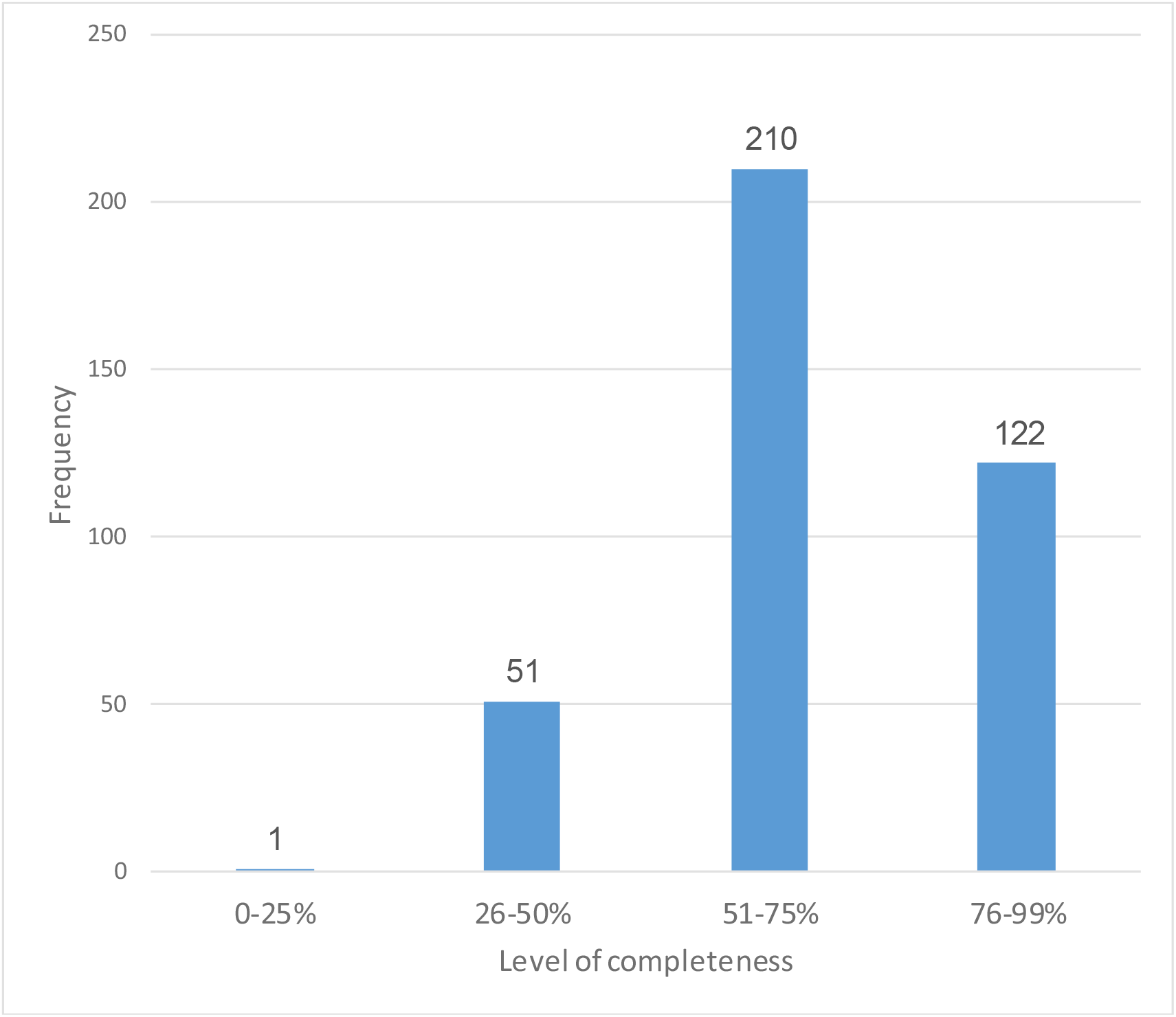
The distribution of referral forms based on its relevant data use on maternal care services (n=384).

The utilization of relevant data among these Referral forms for maternal care services were categorized according to the number of data entries as follows; 0-25% utilization (0-4 entries), 26-50 utilization (5-8 entries), 51-75% utilization (9-12 entries), and 76-99% utilization (13-15 entries). Among this study population, it was found out that there were five (5) form styles received by the receiving facility. It covered varied data but this study identified only those 16 relevant criteria necessary for the continuum of care among referred WRA. Likewise, the current study showed that majority of 384 randomly selected referred WRA (210, 51-75%) had utilized relevant data for maternal care services, and there was no Referral form which utilized the 16 data entries relevant to maternal care services.

## CONCLUSIONS

Local government unit-owned level 1 public hospital, as the receiving facility, received 3,330 referred WRA for 2019. Using the study sample population (384), the study concluded that only 126 referred WRA used the standard Referral form per Department of Health requirement for inter-agency referrals. And based on this standard Referral form, no Referral form was compliant with relative to the to the 14 criteria identified.

Among the top 5 reasons of referring WRA, three were preventable cases and had social and health-seeking behavior implications among the WRA, namely; teenage pregnancy, post-term, and low hemoglobin count. The other two were medically indicated cases as complications to pregnancy such as elevated blood pressure and breech presentation. Furthermore, half of the study population visited the receiving facility late or more than 7 days from the time of referral. Partograph, as a required tool for appropriately managed referred WRA, was attached to eight referral forms. These needs further evaluation and it is beyond the scope of the current study (document analysis only) since partograph are expected to be attached to the patients’ charts for continuity of maternal care.

To improve the Referral form data uptake, this study recommends to include the following data for a continuum of maternal care up to the receiving facility in all Referral forms, namely: 1) last menstrual period, 2) expected date of confinement, 3) age of gestation, 4) fundic height, 5) fetal heartbeat, and WRA 6) body weight, which were used consistently more than 60% during the referral of women of reproductive age. Moreover, revision of the DOH Referral form is highly recommended to all health care givers at the barangay level (other health facilities), standardized, and for regulary monitoring for compliance and monitoring. As an output of the local referral system, this document analysis will have a greater impact to a sustainable and customized data collection. Moreover, the outcome of this functional referral system will highly signifies an improved maternal mortality rate (MMR) and an infant mortality rate (IMR).

## Supporting information

SQUIRE Guidelines

Disclosure of the study

## Data Availability

All data produced in the present study are available upon reasonable request to the author.

## ACKNOWLEDGMENTS

The author would like to acknowledge the management of Justiniano R. Borja General Hospital for allowing to conduct the study and the hospital’s Delivery Room Unit which provided the necessary documents for review. The author would also like to acknowledge the Medical Officers from the City Health Office, and the panel who tirelessly reviewed the manuscript, and the regional office of the Department of Health. There were no financial fundings for this study.

## Appendix I

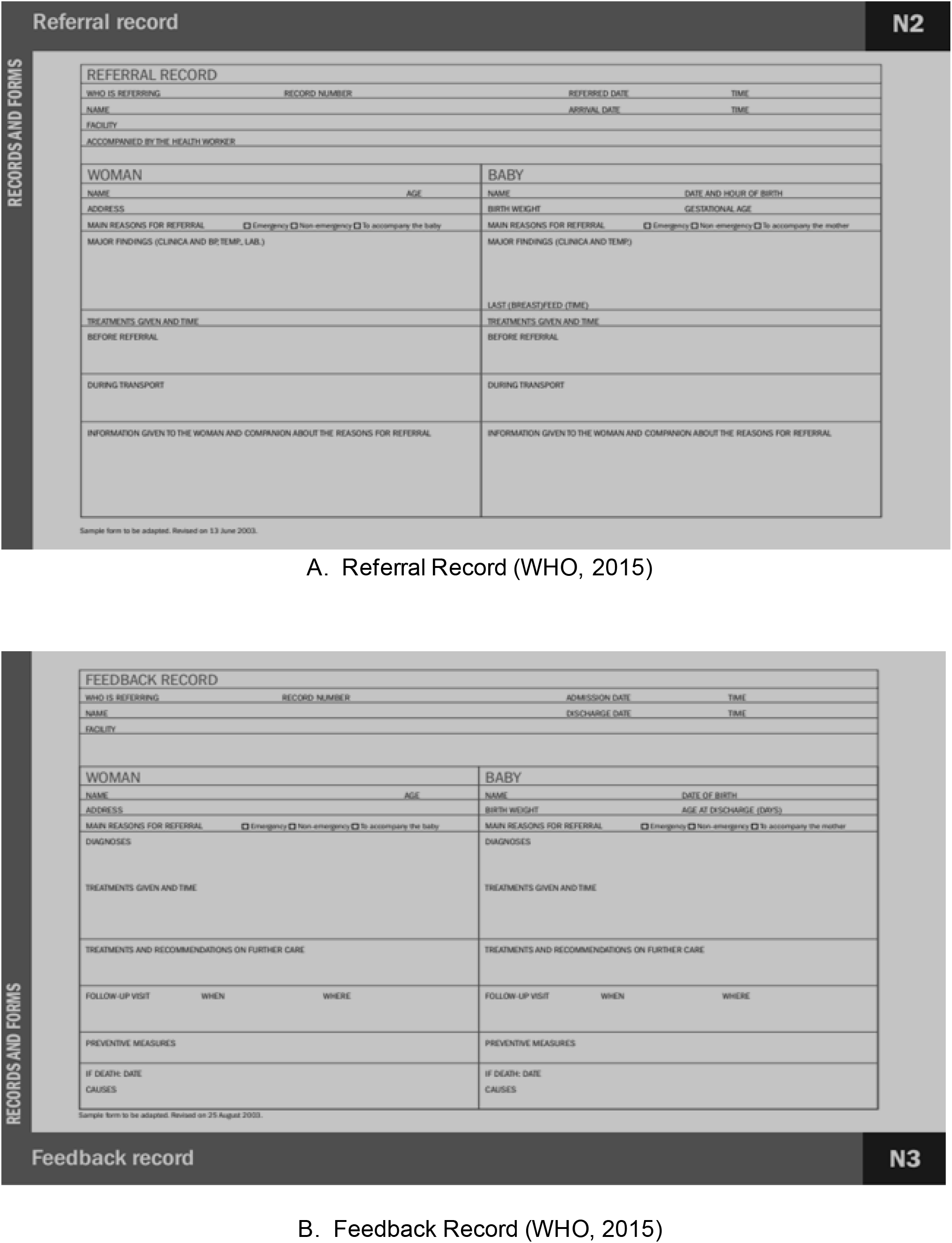

## Appendix II BEmONC/CEmONC* Referral Form

DOH-Northern Mindanao Health System Referral System, 2015 (page 89)

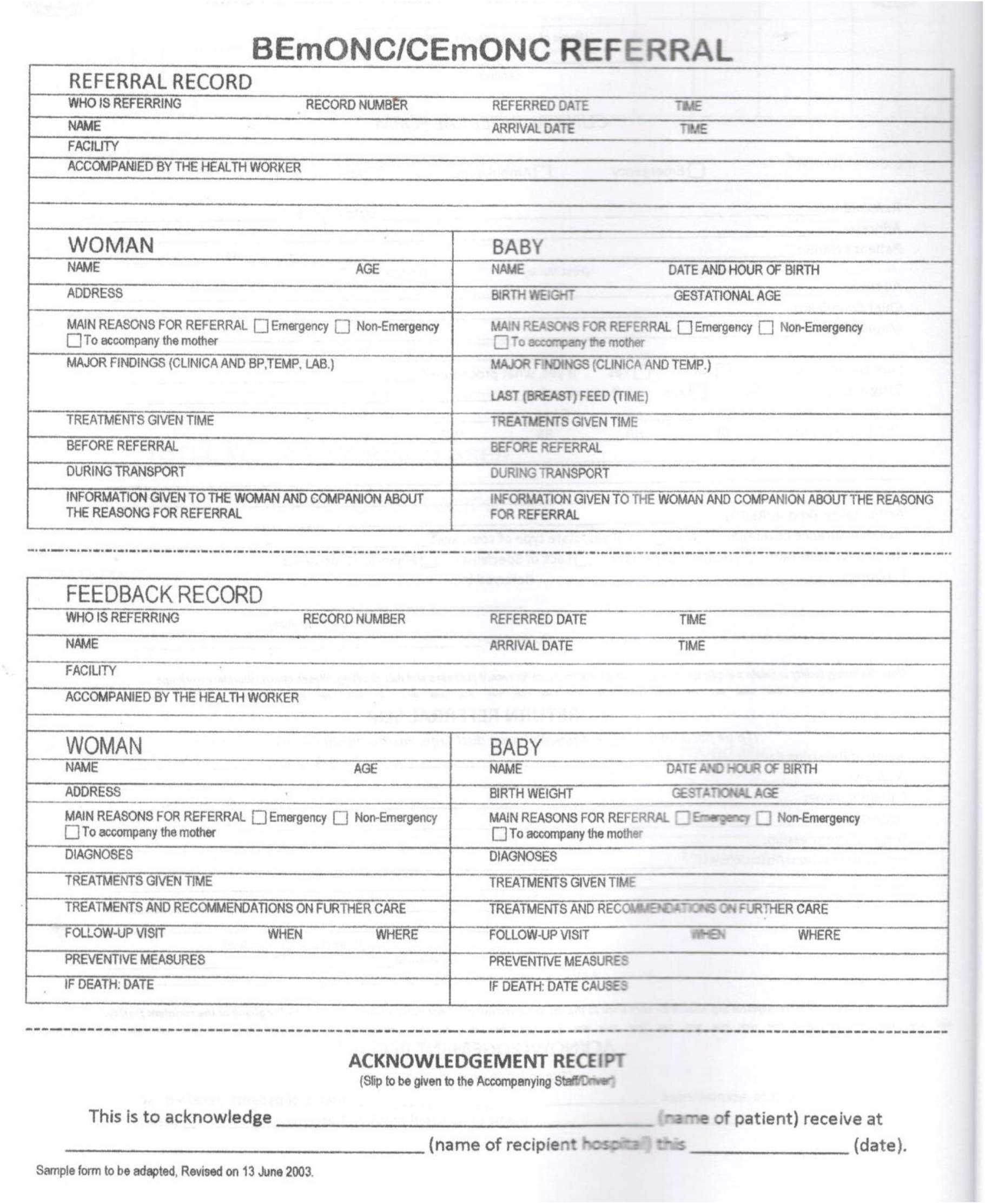

*\*Basic emergency obstetric and Newborn Care (BEmONC)/Comprehensive Emergency Obstetric and Newborn Care (CEmONC)*.

