## Supplementary material for "“Assessment of Referral System on Maternal Services in Cagayan De Oro City”": SQUIRE Guidelines

| | |
|---|---|
| | Referred women of reproductive age benefits from this functional referral system by continuing the initial management and appropriately managed in the next-level of management. Using the Referral form, referral system was assessed by the completeness of the data, and the utilization of the Referral form by each healthcare workers and the referring health facility, and referring back to these referring health facilities. |
| **6.Specific aims** | This study aims to assess the referral system for maternal services in terms of the standard Referral Form, compliance of the health care professionals in using the forms, and the utilization of the data in the referral forms based on 16 criteria for maternal services. |
| **7. Context** | This is a retrospective-document analysis designed to assess the referral system among 3330 randomly selected referred women of reproductive aged (WRA) 15 to 49 years old in Cagayan De Oro City, Philippines from January 2019 to December 2019 in a level 1 public hospital (receiving health facility), The city has 80 barangays with health centers as the referring health facility, referring women of reproductive age for next-level management among high-risk women. There was no actual encounter with this 384-study population which was selected through random sampling of the received referral forms (32 samples per month from January 2019 to December 2019). |
| **8. Interventions** | Received and compiled referral forms were assessed in two ways.<br>First, the received referrals forms were assessed as COMPLIANT if met with the 14/14 criteria. Based on the 14 criteria (compared to the standard Referral form), non-compliant Referral forms were categorized as follows; 0-25% compliant (0-4 entries), 26-50% compliant (5-7 entries), 51-75% compliant (8-10 entries), and 76-99% compliant (11-13 entries). These 13 entries should be filled up were the following: (1) name of the health care provider referring, (2) referring facility, (3) accompanied by the healthcare provider, (4) date and (5) time of referral, (6) age and (7) address, (8) main reason for referral, (9) major findings, (10) treatment given, (11) treatment before and (12) during referral, and (13) information given to the WRA or companion. The presence or a detached (14) Acknowledgement Receipt was the last criteria.<br><br>Secondly, these compiled referral forms used several template and data entries. Thus, this study assessed the relevance of these data relative to maternal services (16/16, 100%). Fifteen (15) criteria using the form and the use of partograph were the variables for maternal services. Likewise, using the 16 criteria for maternal services, Referral forms were categorized accordingly; 0-25% utilized the 16 criteria (0-4 entries), 26-50% utilized (5-8 entries), 51-75% utilized (9-12 entries), and 76-99% utilized (13-15 entries). These 16 variables are necessary for the re-evaluation of WRA especially if in active labor, shortens the history taking, and more focused on their needs. These variables were presented in the Referral Form Assessment Tool (Maternal services), and these are the following: 1) age of the patient, 2) blood pressure monitoring, 3) OB score written as G_P_, 4) reason/s for referral, 5) date of referral, 6) name of referring facility, 7) name of the health care provider from the referring unit, 8) date received, 9) detached acknowledgement receipt, (10) last menstrual cycle (LMP), (11) expected date of confinement (EDC), (12) age of gestation (AOG), (13) fundic height (FH), (14) fetal heart beat (FHB), (15) body weight of WRA (BW), and (16) use of partograph. |
| **9. Study of the interventions** | Appropriate correlations among variables and between the data collected from the referring facilities and receiving facility were analyzed qualitatively. |
| **10. Measures** | The minimum number of criteria to use in assessing a compliant or complete Referral forms were based on the Referral System Assessment (RSA) Instrument developed by USAID, PEPFAR, and MEASURE Evaluation (2013). Two separate focus-group discussions among healthcare providers were conducted to determine and verify the identified gaps on referral system. Likewise, key-informant interviews (KII) were conducted among program managers of maternal services from the City Health Office. The content of KII discussions followed the revised RSA Tool for KII and FGD based on the Guide in Conducting FGD and KII.<br><br>***Definition of Terms.***<br>1.Compliant Referal Form. Refers to the availability of 14 data entries filled up in the Referral form and detached Acknowledgement Receipt based on the standard Referral form (14/14, 100%). Moreover, it was based on the following 14 criteria: (1) name of the health care provider referring, (2) referring facility, (3) accompanied by the healthcare provider, (4) date and (5) time of referral, |

| | |
|---|---|
| | standard Referral form per Department of Health requirement for inter-agency referrals. And based on this standard Referral form, no Referral form was compliant with relative to the to the 14 criteria identified.<br>Among the top 5 reasons of referring WRA, three were preventable cases and had social and health-seeking behavior implications among the WRA, namely; teenage pregnancy, post-term, and low hemoglobin count. The other two were medically indicated cases as complications to pregnancy such as elevated blood pressure and breech presentation. Furthermore, half of the study population visited the receiving facility late or more than 7 days from the time of referral. Partograph, as a required tool for appropriately managed referred WRA, was attached to eight referral forms. These needs further evaluation and it is beyond the scope of the current study (document analysis only) since partograph are expected to be attached to the patients' charts for continuity of maternal care. |
| **15. Interpretation** | The currently used referral forms by the healthcare professionals came in different forms. The healthcare professionals were not compliant (14 criteria) in utilizing the data entries of the recommended template. Likewise, data entries relevant to the continuum of maternal services (16 criteria) need to be included in the current referral forms. |
| **16. Limitations** | The study population were limited to the women of reproductive aged (WRA, 15– 49 years old) who were referred from their respective barangay health centers (referring facility) of Cagayan De Oro City to avail higher level of maternal services. Referral forms were retrieved and data collection were gathered retrospectively based on these forms. There were no interactions nor data verification with the teenage and women of reproductive age. Referral forms were randomly selected among the compiled forms at JRBGH from January 2019 to December 2019. Referred WRA due to gynecologic cases, non-residents of Cagayan De Oro City, resident of the city but referred from other health facilities outside of Cagayan De Oro City, and use of prescription form as Referral form were excluded from the study. |
| **17. Conclusions** | To improve the Referral form data uptake, this study recommends to include the following data for a continuum of maternal care up to the receiving facility in all Referral forms, namely: 1) last menstrual period, 2) expected date of confinement, 3) age of gestation, 4) fundic height, 5) fetal heartbeat, and WRA 6) body weight, which were used consistently more than 60% during the referral of women of reproductive age. Moreover, revision of the DOH Referral form is highly recommended to all healthcare givers at the barangay level (other health facilities), standardized, and for regular monitoring for compliance and evaluation. As an output of the local referral system, this document analysis will have a greater impact to a sustainable and customized data collection. Moreover, the outcome of this functional referral system will highly signify an improved maternal mortality rate (MMR) and an infant mortality rate (IMR). |
| **Other Information** | |
| **18. Funding** | There was no funding that support this work. |
